# Association between asthma and suicidality in 9-11-year-old children

**DOI:** 10.1101/2021.10.23.21265416

**Authors:** Kevin W. Hoffman, Elina Visoki, Stirling T. Argabright, Grace E. Didomenico, Barbara H. Chaiyachati, Tyler M. Moore, Ran Barzilay

## Abstract

**Background:** Suicidal thoughts and behavior (STB) in children are a growing health concern, and more data is needed regarding their biological underpinnings. Immune processes such as inflammation have been associated with STB, primarily in adults. Asthma is a common chronic inflammatory disorder in children and has been associated with STB in adolescent and adult populations, but data in children is lacking. We wished to study associations of asthma with childhood STB given asthma’s potential as a clinically relevant model for childhood chronic immune dysregulation.

**Methods:** Using data from the Adolescent Brain Cognitive Development (ABCD) Study (N=11,878, 52% males, mean age 9.9 years at baseline assessment and 10.9 years at 1-year follow up), we assessed associations between asthma and STB at both baseline and 1-year follow up.

**Results:** We found that asthma at baseline assessment (n=2,214, 18.6%) is associated with STB, controlling for multiple confounders including demographics, socioeconomic factors and environmental confounders such as air pollution (odds ratio (OR)=1.2, 95%CI 1.01-1.42, P=0.039). Indicators of recently active asthma were not significantly associated with suicidality at baseline assessment (currently taking asthma medication: OR=1.22, 95%CI 0.93-1.60, P=0.146), or at 1-year follow up (past year asthma-related clinical visit: OR=1.13, 95%CI 0.87-1.47, P=0.357). Proxy-measures of asthma severity (number of asthma medications or clinical visits) did not reveal a significant dose response relationship with STB.

**Conclusions:** Findings suggest an association between history of asthma and STB in children, which may not be related to asthma disease state. Further research is needed to investigate mechanisms underlying this relationship.

## Introduction

Compromised physical health and inflammatory dysregulation are associated with suicidal behaviors (1–3). Children with asthma commonly possess both of these traits. Asthma has been independently associated with suicide risk in adolescent and adult populations (4), but more data is needed in children and early adolescents. Considering that asthma is the most common chronic disease in children, with steadily increasing rates, understanding suicide risk in this population is critical (5). Compounding this urgency is a steadily increasing rate of suicidal thoughts and behaviors (STB) in children aged 10-14 years (6–9).

While asthma itself is associated with STB risk, it is frequently controlled with medications that should also be assessed for STB risk. Recently, the asthma medication Montelukast was required by the FDA to carry a boxed warning for serious mental health side effects including suicidal thoughts or actions following observational study of 15 years of use (10). Other studies have additionally linked asthma medications with suicide risk in adults (11). Similar associations have been reported between immune modulating drugs and STB in adults (11,12).

The purpose of this study was to evaluate the association of asthma and asthma medication use with STB in 9-11-year-old children. We hypothesized that asthma, as an example of an immune dysregulation illness, would be associated with childhood STB. We used data obtained from the Adolescent Brain Cognitive Development (ABCD) Study (13), a large, diverse youth sample (N>11,000) with extensive phenotyping for sociodemographic factors, psychopathology, medical conditions and treatments, and multiple environmental exposures (14).

## Methods and Materials

### Study design

We employed a cross-sectional design examining associations between history of asthma or current use of asthma medications and STB at baseline ABCD Study assessment, and the association between past-year asthma/asthma attack and STB in 1-year follow-up assessment.

### Sample

The sample is composed of all ABCD Study participants who completed the baseline assessment (N=11,878) or the 1-year follow-up assessment (N=11,235). The ABCD Study sample includes a cohort of children aged 9–10 years at baseline, recruited through school systems (15). Participants were enrolled at 21 sites, with catchment area encompassing over 20% of the entire US population in this age group. We included data from the ABCD Study data release 3.0 (https://abcdstudy.org/). All participants gave assent. Parents/caregivers signed informed consent. The ABCD Study protocol was approved by the University of California, San Diego Institutional Review Board (IRB), and was exempted from a full review by University of Pennsylvania IRB.

### Exposures

A history of asthma was considered for all participants that endorsed at least one of the following items at baseline assessment: parent report of asthma diagnosis at baseline assessment (referred throughout the manuscript as *parent report*, variable medhx_2a from the ABCD Summary Scores Medical History); parent report of asthma attack for which the participant was seen by a healthcare provider (referred throughout the manuscript as *asthma attack*, variable medhx_6l from the ABCD Summary Scores Medical History); and parent report of child taking asthma medications in the past 2 weeks (referred throughout as *asthma medications*, also assessed at baseline, data from the ABCD Parent Medications Survey Inventory). We created a composite measure of these measures referred throughout the manuscript as *asthma composite*, or simply, *asthma*.

*Recently active asthma* was considered for all participants that endorsed at least one of the following items at the 1-year follow-up assessment: having been to the doctor since the baseline assessment for asthma (variable medhx_2a_l) or for asthma attack (variable medhx_6l_l).

Asthma medications taken in the past 2 weeks (binary variable) as reported at baseline ABCD Study assessment included: beta-agonists, inhaled corticosteroids, leukotriene modifiers, and others (see **Supplementary Table 1** for a full list of asthma medications).

Asthma severity was estimated by two variables available at the baseline assessment: a) The number of parent-reported clinical visits due to asthma attacks (medhx_ss_6l_times_p, continuous variable); and b) The number of asthma medications that the participant was taking (continuous variable).

### Outcome measures

The ABCD Study assessed suicidal ideation and attempt (past or current) as part of the validated and computerized Kiddie-Structured Assessment for Affective Disorders and Schizophrenia for DSM-5 (KSADS-5) (16). Items relating to self-injurious behavior without suicidal intent were not included in the current analysis. As the proportion of suicide attempts was low, and to mitigate risk of type I error caused by multiple testing, we combined suicidal ideation and attempt, consistent with previous analyses (17,18). Thus, STB was defined as a single binary measure termed “suicidality,” considered for children who endorsed past/present suicidal ideation (SI) and/or suicide attempt (SA) at ABCD Study baseline or 1-year follow-up assessment. Prevalence of SI and SA is detailed in **Supplementary Table 2**. As prior studies (including in the ABCD Study) showed poor youth-caregiver agreement on STB (18–20), we focused on child-report in the current analysis.

### Covariates

Covariates were obtained from ABCD Study baseline data and included demographics: age, sex, race (Black, White, Other), and Hispanic ethnicity (used in Model 1 described below); household-level socioeconomic status (SES) variables: household income, average parent education, and maternal age (added in Model 2 described below); and neighborhood-level factors: area deprivation index, population density, NO2 and PM 2.5 (annual average at 10×10km^2^), and proximity to major roads (added in Model 3 described below).

### Statistical Analysis

We used the SPSS 26.0 statistical package and R for our data analyses. Mean (standard deviation [SD]) and frequency (%) were reported for descriptive purposes. Univariate comparisons were made using t-tests or chi-square tests, as appropriate. We employed listwise deletion for participants with missing data, the rate of which was lower than 2.3% for all variables except neighborhood-level variables (5.9%) and household income (8.6%).

The analytic plan and hypotheses were preregistered on Open Science Framework (https://osf.io/nxeys/) in July 2021 and analyses were conducted in August-September 2021. Data preprocessing and analysis are detailed at https://github.com/barzilab1/ABCD_asthma_inflammation_k.

### Main Analyses

We tested a set of binary logistic regression models with asthma exposures as independent variables (IV) and suicidality as the dependent variable (DV). We first tested models using the baseline data with asthma composite and each of its comprising items (parent report, asthma attack and asthma medications) as IVs, and suicidality as DV. Then, we tested a cross-sectional model using the 1-year follow-up data with currently active asthma as IV and suicidality as DV.

In all the above-described models, we added covariates in a stepwise manner, beginning with demographics (Model 1), then adding household-level SES (Model 2), and then adding neighborhood-level exposures (Model 3).

### Sensitivity Analyses

We ran two sensitivity analyses to address the specificity of our findings from the main analyses: to address specificity of the exposure (asthma), we ran similar models as above using the measure “ever seen a doctor for broken bones” (measure medhx_6a in ABCD Study) as the IV; to address the possibility that severity of asthma affects the associations identified in the main analyses, we tested models using proxies of asthma severity (“number of asthma attacks” or “number of asthma medications”) as the IV.

## Results

### Prevalence of Asthma and STB in ABCD

Among 11,878 children at baseline ABCD Study assessment (mean age=9.9 years [SD=0.6], 6,196 boys [52.2%]), 2,214 (18.6%) endorsed at least one asthma exposure and met our definition as having a history of asthma (asthma composite). Of which, 2,039 (17.2%) were described by their caregivers as having asthma, 809 (6.8%) had been seen by a medical provider for an asthma attack, and 698 (5.9%) were taking asthma medications. Overall, 1,040 (8.8%) reported suicidality. Demographic and clinical characteristics of the baseline sample are detailed in **Table 1**.

**Table 1:** Sample socio-demographics, asthma, and STB prevalence in the ABCD study.

|  |  |
| --- | --- |
| **Total sample (baseline)<sup>a</sup>** | **N=11,878** |
|  | **Mean (SD)** |
| **Age at baseline assessment, years** | 9.92 (0.62) |
|  | **n (%)** |
| **Sex, male** | 6,196 (52.2%) |
| **White** | 8,805 (74.1%) |
| **Black** | 2,518 (21.2%) |
| **Asian** | 752 (6.3%) |
| **Hispanic** | 2,411 (20.3%) |
| **Asthma variables** |  |
| **Parent report of asthma** | 2,039 (17.2%) |
| **Asthma attack** | 809 (6.8%) |
| **Asthma medications** | 698 (5.9%) |
| **Asthma composite<sup>b</sup>** | 2,214 (18.6%) |
| **Recently active asthma<sup>c</sup>** | 891 (7.5%) |
| **Asthma medication variables** |  |
| **Non-steroids (bronchodilators, leukotriene modifiers)** | 697 (5.9%) |
| **Steroids** | 46 (0.4%) |
| **Suicidal thoughts and behavior (STB) measures** |  |
| **Ever suicidal ideation (SI)** | 1,025 (8.6%) |
| **Ever suicide attempt (SA)** | 156 (1.3%) |
| **Suicidality (ever SI or SA)** | 1,040 (8.8%) |

<sup>a</sup>For all variables, missing data rate was less than 2.3% (273 participants out of the 11,878 participants at baseline ABCD assessment).
<sup>b</sup> A history of asthma was considered for all participants that endorsed at least one of the following items at baseline assessment: parent report of asthma diagnosis at baseline assessment; parent report of asthma attack for which the participant was seen by a healthcare provider; and parent report of child taking asthma medications in the past 2 weeks. <sup>c</sup> Recently active asthma was considered for all participants that endorsed at least one of the following items at the 1-year follow-up assessment: having been to the doctor since the baseline assessment for asthma or for asthma attack.

### Association of asthma with suicidality in baseline ABCD Study assessment

Univariate comparison revealed that children with a history of asthma (asthma composite) endorsed suicidality at higher rates than children without asthma (**Table 2**), with 10.3% rate endorsement of suicidality among participants with asthma compared to 8.5% among the non-asthma participants (Pearson chi-square (df=1)=7.119; P=0.008). In contrast, there was no difference in rates of suicidality between participants who had ever broken bones (n=1,706, of which 9.4% endorsed suicidality) and those who had not (n= 10,168, of those 8.7% endorsed suicidality, P = 0.342).

**Table 2:**
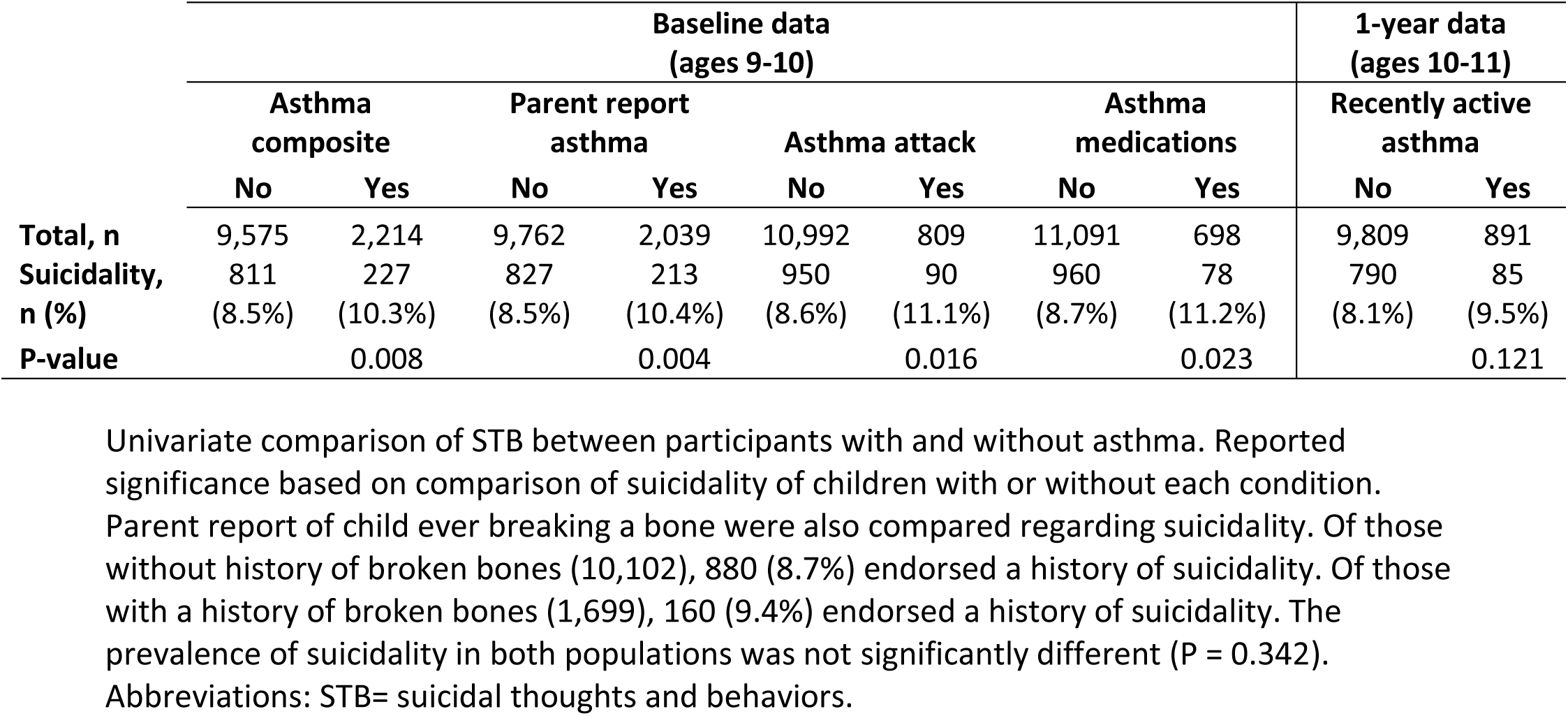
Univariate comparison of STB between participants with and without asthma.

To address the specificity of the above association we conducted multivariable analysis to account for potential confounding effects (**Table 3**). The association between asthma and suicidality was not statistically significant when co-varying for demographic factors (Model 1; odds ratio [OR]=1.17, 95% confidence interval [CI] 0.99-1.37, P=0.06), but when we co-varied for household SES (measured by income, average parent education, and maternal age), the association was significant (Model 2; OR=1.2, 95%CI 1.01-1.42, P=0.035). The asthma-suicidality association remained significant when including neighborhood environment confounders including area deprivation index, population density, air pollution (NO2, PM 2.5), and proximity to major roads (Model 3; OR=1.2, 95%CI 1.01-1.42, P=0.039). In contrast to asthma, a history of having broken a bone did not show an association with suicidality in any of the multivariable models (all P’s>0.279).

**Table 3:**
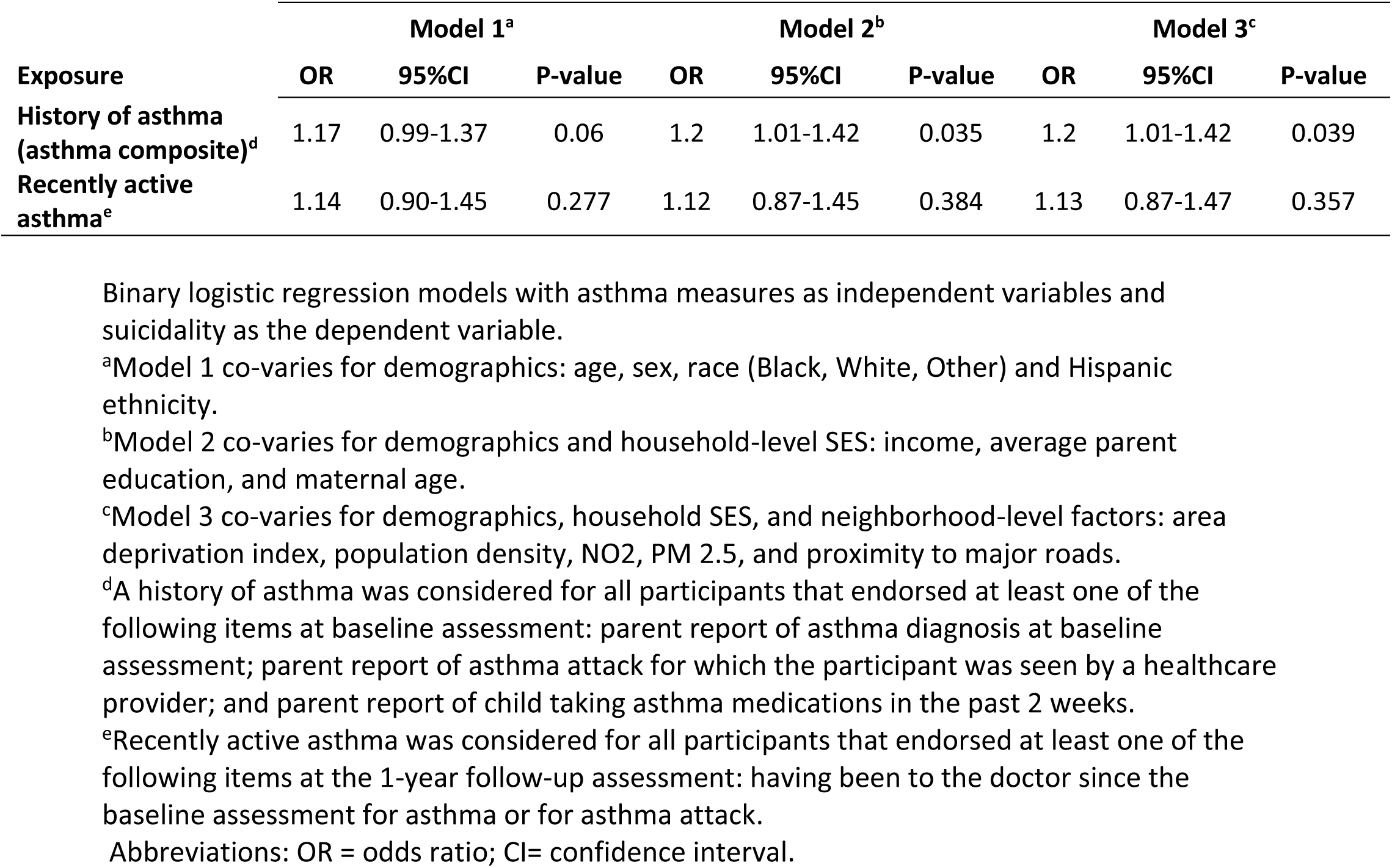
Association between asthma and suicidality in the ABCD Study.

When using different definitions of asthma exposures (that comprise the asthma composite variable), multivariable models showed statistically significant associations with suicidality for the exposures of parent report of asthma and having seen a healthcare provider for asthma attacks, whereas none of the multivariable models showed a significant association between taking asthma medications and suicidality (**Supplementary Table 3**).

### Association of having recently active asthma with suicidality in the 1-year follow-up ABCD Study assessment

Among participants of the 1-year follow-up ABCD Study assessment (mean age 10.9), no significant difference was observed comparing suicidality rates among participants having recently active asthma (estimated by going to see a doctor for asthma/asthma attack in the time between baseline and 1-year follow up ABCD Study assessment) compared to their counterparts (9.5% Vs 8.1%, P=0.121, **Table 2**). Multivariable analyses revealed no association of recently active asthma with suicidality (**Table 3;** when co-varying for demographics, household SES and neighborhood factors: OR=1.13, 95%CI 0.0.87-1.47, P=0.357).

### Association of asthma severity proxies with suicidality

To explore a possible dose relationship between asthma severity and suicidality, we evaluated two metrics of asthma severity proxies that are available in the ABCD Study baseline data: reported number of asthma attacks and total number of asthma medications. While both measures showed a direction of association that might suggest a dose response, there was not a statistically significant association in any of the tested models (in models that co-varied for demographics, household SES and neighborhood factors: OR for each additional clinical visit due to asthma attack=1.04, 95%CI 0.99-1.1, P=0.113; OR for each additional asthma medication=1.11, 95%CI 0.95-1.29, P=0.203, see **Supplementary Table 4** for full models’ statistics).

## Discussion

We describe an association between history of asthma and suicidality in a large, diverse sample of preadolescent US children (age 9-10). The association was significant when accounting for multiple SES household variables and neighborhood confounders. Furthermore, sensitivity analyses showing a lack of association with suicidality of a non-inflammatory medical condition (bone fractures), further supports the specificity of findings. These results add to existing literature on the associations between asthma and suicidality in adolescents (21–25) and adults (26–35), and support the biological evidence for an association between immune dysregulation and suicidality (36–38), presenting early in the lifespan. Notably, while suicidality itself is an important target for investigation in terms of its association with immune dysregulation (39), it may also be a key non-specific indicator of significant psychological distress in young children. Indeed, early life suicidality is strongly tied to stress exposure (40–42), which in turn is associated with immune dysregulation (43,44). Our findings therefore provide a clinical indication for the link between inflammation (i.e., asthma) and suicidality, and support the notion that immune mechanisms may be involved in the development of suicidality.

While asthma is common, it is not evenly distributed across the population, with different prevalence between genders, racial groups, SES, and populations from urban vs rural settings (45,46). Similarly, gender, race, and SES, all associated with various levels of discrimination, are all associated with youth suicidality (47–51). A major strength of the ABCD Study is that its large sample size and deep phenotyping allows sufficient power to account for all the above potential confounders and support the specificity of our findings showing a positive association of asthma and childhood suicidality. Notably, while this association remained significant for all three multivariable models for asthma identified by parent report (demographic in Model 1; household SES in Model 2; neighborhood environment in Model 3), the exposure of taking asthma medications did not show significant association with suicidality in any of the models, with substantial decrease in statistical significance once SES was included in the model (P = 0.069 for Model 1, P = 0.123, 0.126 for Models 2 and 3 respectively). One potential explanation for this finding is that inadequate access to asthma medication in children from lower-resourced SES backgrounds may be confounding the association between taking asthma medication and suicidality.

In addition to the asthma-suicidality association in baseline data at ages 9-10, we used the 1-year follow up ABCD Study data to attempt to assess whether recently active asthma (seeing a doctor for asthma or asthma attack in the past year) is associated with suicidality, and found no evidence to support such association. Furthermore, the fact that in the baseline data, there was no association between currently taking asthma medication and suicidality may also indicate a lack of association between the recency of active asthma and suicidality. Together these findings might suggest that it is a history of asthma (i.e., “trait”), rather than active asthma (i.e., “state”), that drives the association with suicidality. It is possible that underlying asthma factors, such as chronic immune dysregulation, and not factors relating to asthma attacks themselves (e.g., psychological distress from asthma symptoms) explain the association observed between asthma and suicidality. In this case, the state of having asthma would be expected to be associated with suicidality, but proximity to an asthma attack (and consequently the increased likelihood of taking asthma medications) would not necessarily be associated with suicidality, especially as the power to detect these effects diminishes when narrowing on a population with recently active disease (n=891 with recently active asthma compared to n=2,214 with a history of asthma in the current study).

As a final aim, we wished to investigate possible dose relationships between asthma and suicidality. Establishing a dosage effect helps build an argument for a causal relationship between two correlated factors and can potentially shed light on mechanisms through which asthma may influence suicidality. The two severity metrics we chose (lifetime number of asthma attacks and total number of asthma medications) were chosen because they provided both medication-independent and -dependent measures of severity, which provides a means of distinguishing between suicide risk driven by asthma vs suicide risk driven by asthma medications. While neither metric showed statistical significance, both correlations showed a positive direction, suggesting that our study population may have been underpowered to address these questions. It is also possible that we were looking at the wrong dose relationship. If the correlation between asthma and suicidality is primarily driven by inflammatory dysregulation or common genetic linkages, it may be the case that the number of asthma attacks or asthma medications is irrelevant to suicide risk and instead inflammatory or genetic factors may alternatively establish causality.

This study suggests a clinical association between asthma and suicidality in children and should propel future research aiming to identify mechanisms that drive this association. A few mechanisms have been previously proposed to explain the connection between asthma and suicidality (4). First, asthma symptoms may cause psychological distress consequently leading to suicidality. Second, asthma’s immune dysregulation may drive increased depression (52,53), which in turn drives increased suicidality. Third, there may be common genetic linkages between asthma and mood. While originally proposed to explain a connection between asthma and depression, this framework is useful for suicidality as well, with the added consideration that asthma medications themselves may drive suicidality, as evidenced by growing concern surrounding Montelukast (54).

Given these proposed mechanisms and the results of our study, several avenues of investigation present themselves. Future studies that include medical records of children with asthma may allow for greater power to assess associations between asthma medications and suicidality, and in-depth psychological assessment of the subset endorsing suicidality may further allow investigation between specific asthma symptoms and suicidality. Additionally, if, as we hypothesize, immune dysregulation is important in the link between asthma and suicide, similar associations would be expected in children subject to other immune dysregulatory and pro-inflammatory processes. Finally, our results lay the groundwork for future genetic studies investigating linkages between asthma and suicidality in youth.

## Limitations

Our findings should be viewed in light of several limitations. First, the ABCD Study was not intended specifically to address this study’s questions, hence the phenotyping for asthma and its severity was established without a clinician’s rating and/or specific asthma scales, and we relied instead on parent report of asthma diagnosis, report of asthma attacks, and asthma medications. Another limitation is the cross-sectional nature of this study. More studies are needed to investigate longitudinal relationships between asthma and suicidality in the ABCD Study and other cohorts. Additionally, the current study does not include biological measures of immune dysregulation, which should be explored in future works as ABCD Study releases biological specimens that can allow immune phenotyping. Lastly, while the ABCD Study is large, including 21 sites across the US, our findings need to be generalized and replicated in other youth datasets.

## Conclusions

This study describes an association between history and asthma and suicidality in a large community sample of preadolescent children. The large number of children studied, the within-sample diversity, and the broad nature of data collected allowed controlling for multiple confounders, providing a basis to investigate mechanisms underpinning this asthma-suicidality association.

## Data Availability

Qualified researchers can request access to ABCD shared data through the NIH data archive.

https://nda.nih.gov/abcd/request-access

## Acknowledgement

Data used in the preparation of this article were obtained from the Adolescent Brain Cognitive DevelopmentSM (ABCD) Study (https://abcdstudy.org), held in the NIMH Data Archive (NDA). This is a multisite, longitudinal study designed to recruit more than 10,000 children age 9-10 and follow them over 10 years into early adulthood. The ABCD Study® is supported by the National Institutes of Health and additional federal partners under award numbers U01DA041048, U01DA050989, U01DA051016, U01DA041022, U01DA051018, U01DA051037, U01DA050987, U01DA041174, U01DA041106, U01DA041117, U01DA041028, U01DA041134, U01DA050988, U01DA051039, U01DA041156, U01DA041025, U01DA041120, U01DA051038, U01DA041148, U01DA041093, U01DA041089, U24DA041123, U24DA041147. A full list of supporters is available at https://abcdstudy.org/federal-partners.html. A listing of participating sites and a complete listing of the study investigators can be found at https://abcdstudy.org/consortium_members/. ABCD consortium investigators designed and implemented the study and/or provided data but did not necessarily participate in analysis or writing of this report. This manuscript reflects the views of the authors and may not reflect the opinions or views of the NIH or ABCD consortium investigators.

